# Evaluating a Genome-Wide Polygenic Score for Handgrip Strength and Its Interplay with Leisure-Time Physical Activity Across the IGEMS Twin Cohorts

**DOI:** 10.1101/2025.08.07.25333195

**Authors:** Päivi Herranen, Teemu Palviainen, Marianne Nygaard, Ida K Karlsson, Anbupalam Thalamuthu, Karen A Mather, Chandra A. Reynolds, Matthew S. Panizzon, Taina Rantanen, Deborah Finkel, Margaret Gatz, Nancy L. Pedersen, IGEMS Consortium, Jaakko Kaprio, Elina Sillanpää

## Abstract

**Background**

Polygenic scores (PGSs) may help assess genetic predisposition to multifactorial traits. We examined whether age, sex, and leisure-time physical activity (LTPA) modify the association between a PGS for handgrip strength (HGS) and measured HGS in older adults.

**Methods**

PGS HGS, based on Pan-UK Biobank GWAS data, was calculated for 5103 participants (aged 40–96; 44% women) from eight twin cohorts in Denmark, Sweden, Australia, the United States, and Finland within the IGEMS consortium. Sex-standardized HGS and self-reported LTPA were assessed cross-sectionally. Linear mixed models estimated associations between PGS and HGS, including interactions with age, country, and LTPA, as well as an association between PGS and LTPA. Fixed-effect within-pair models were conducted to assess environmental contributions.

**Results**

Higher PGS was associated with greater HGS (β = 2.14, SE = 0.15, *p* < 0.001), explaining 4.6% of HGS variance overall, with modest variation across countries. In sex-stratified models, PGS explained 5.2% of the variance in females and 4.3% in males. A significant PGS × LTPA interaction (β = –0.034, *p* = 0.013) indicated that LTPA had a stronger effect among individuals with lower PGS HGS. No statistically significant interaction with age was found. The within-pair models offered limited support for the environmental impact of LTPA.

**Conclusions**

PGS for HGS was associated with measured HGS, with effect modification by LTPA. Findings provide some evidence that physical activity may buffer against genetic predisposition to lower HGS. The results highlight the potential of PGSs to capture individual differences in strength-related traits across populations.

## Introduction

The aging process is characterized by an involuntary reduction in muscle mass and strength, resulting from a combination of genetic, environmental, and lifestyle factors, along with their intricate interplay (1). Muscle strength begins to gradually decline from around the age of 30, but significant changes in the aging process occur after the age of 50, with an annual diminishing of muscle strength by 1–2% (2). Low muscle strength is associated with an increased risk of chronic diseases, disability, and mortality (1,3). Among modifiable factors, physical activity (PA) plays a key role in preserving muscle strength and mitigating age-related decline. Regular PA can counteract losses in muscle mass and function, supporting mobility and reducing the risk of disability (3). At the same time, reduced muscle strength may lead to lower PA engagement, creating a feedback loop that accelerates physical decline (4). Conversely, greater muscle strength can enhance one’s ability and motivation to remain active, reinforcing long-term physical function and well-being (5).

Handgrip strength (HGS) is a widely used, reliable marker of overall muscle strength and an important indicator of functional status and health in aging populations (6). HGS, even when measured in midlife, predicts future adverse outcomes, including disability and mortality (7–9). HGS can therefore be considered to signal the individual’s intrinsic physiological capacity to resist functional decline into critical disease and disability levels (10). HGS is a complex, multifactorial trait affected also by internal factors such as age and sex. Males typically exhibit higher HGS throughout the lifespan, and subtle sex differences have been observed in the rate of age-related decline (11,12). Classical twin designs among monozygotic (MZ) and dizygotic (DZ) twins, who share 100% and ∼50% of their segregating genes, respectively, have shown that HGS is moderately to highly heritable, with genetic factors accounting for 30–65% of its variance (13). This heritability appears relatively stable with age (11,12,14), though some findings suggest environmental influences may increase over time (13,15). Evidence regarding sex differences in the genetic architecture of HGS is mixed. While one study reports comparable genetic and environmental contributions across sexes (12), others have found higher heritability in males (11,16) and greater shared environmental influence in females (11). Additionally, the types of environmental factors that contribute to HGS also appear to differ between sexes across adulthood (17).

From a genetic perspective, HGS is a polygenic trait, that is, it is influenced by many genes across different loci, each with a low effect (18,19). A polygenic score (PGS) is a genetic tool used to estimate an individual’s predisposition to a trait or condition, based on the aggregated effect sizes of multiple genetic variants, typically single-nucleotide polymorphisms (SNPs), across the genome (20). While the observed sex differences in the genetic architecture of human traits are generally considered to have limited consequences (21,22), studies have shown that the effects of many disease-specific PGS can vary by age and sex (23). We have recently constructed PGS for HGS and found it to be a reliable predictor of overall muscle strength among older Finnish females, explaining 6.1% of the variation in the measured HGS and 5.4% of the variation in the knee extension strength (24). In this study, we aimed to extend the evaluation of the PGS HGS to international twin cohorts and investigate whether the association between genetic predisposition to HGS and measured strength is modified by age, sex, country of residence, or leisure-time PA (LTPA). We further examined the role of LTPA in modifying genetic effects and assessed its independent contribution using a within-twin pair analysis (25), which effectively controls for both genetic and nongenetic factors that are common to both twins in a pair.

## Method

### Study design and participants

The PGS for HGS was derived using publicly available summary statistics from the Pan-UK Biobank (Pan-UKBB) genome-wide association study (GWAS) of maximum HGS, which included 418 827 individuals aged 40 to 69 years (https://pan.ukbb.broadinstitute.org.2020). We calculated the PGS for HGS in participants of the Interplay of Genes and Environment across Multiple Studies (IGEMS) consortium (26), and first validated it by analyzing its association with measured HGS. We also assessed the extent of missing heritability—defined as the proportion of genetic influence not captured by the PGS—by comparing SNP-based and pedigree-based heritability estimates. We then explored potential effect modification by age, country of residence, and LTPA, and further applied a co-twin control design using within-pair comparisons. The study flow is shown in Figure 1. IGEMS is an international consortium of 20 twin studies focusing on longitudinal twin investigation of adulthood and aging. Our analytic sample comprised 5103 twin individuals drawn from eight independent cohorts, spanning five countries: Denmark, Sweden, Australia, the United States, and Finland. All of them had available PGS, ancestry-informative genetic principal components (PCs) (27), and at least one valid HGS measurement (Supplementary eFigure 1). Of those, 4451 participants had data for both LTPA and body mass index (BMI). A subsample of 1982 complete twin pairs (43% MZ and 57% DZ pairs) was used in within-pair analyses (Supplementary eFigure 2). The specific methods to determine zygosity varied among studies, but in most cases, standard questionnaires were administered, and DNA analysis was conducted to clarify uncertain instances (28). The North West Multi-centre Research Ethics Committee approved the UK Biobank study (21/NW/0157). The IGEMS Consortium was approved by the University of Southern California Institutional Review Board (approval number UP-16-00315). The FITSA data collection was approved by the Ethics Committee of the Central Hospital District of Central Finland (KSSHP 24/2000). All studies obtained informed written consent from all participants and adhered to the principles of the Declaration of Helsinki. Additional cohort-specific details are provided in the Supplementary Materials under *Overview of IGEMS Cohorts Included in the Analysis*.

**Figure 1.**
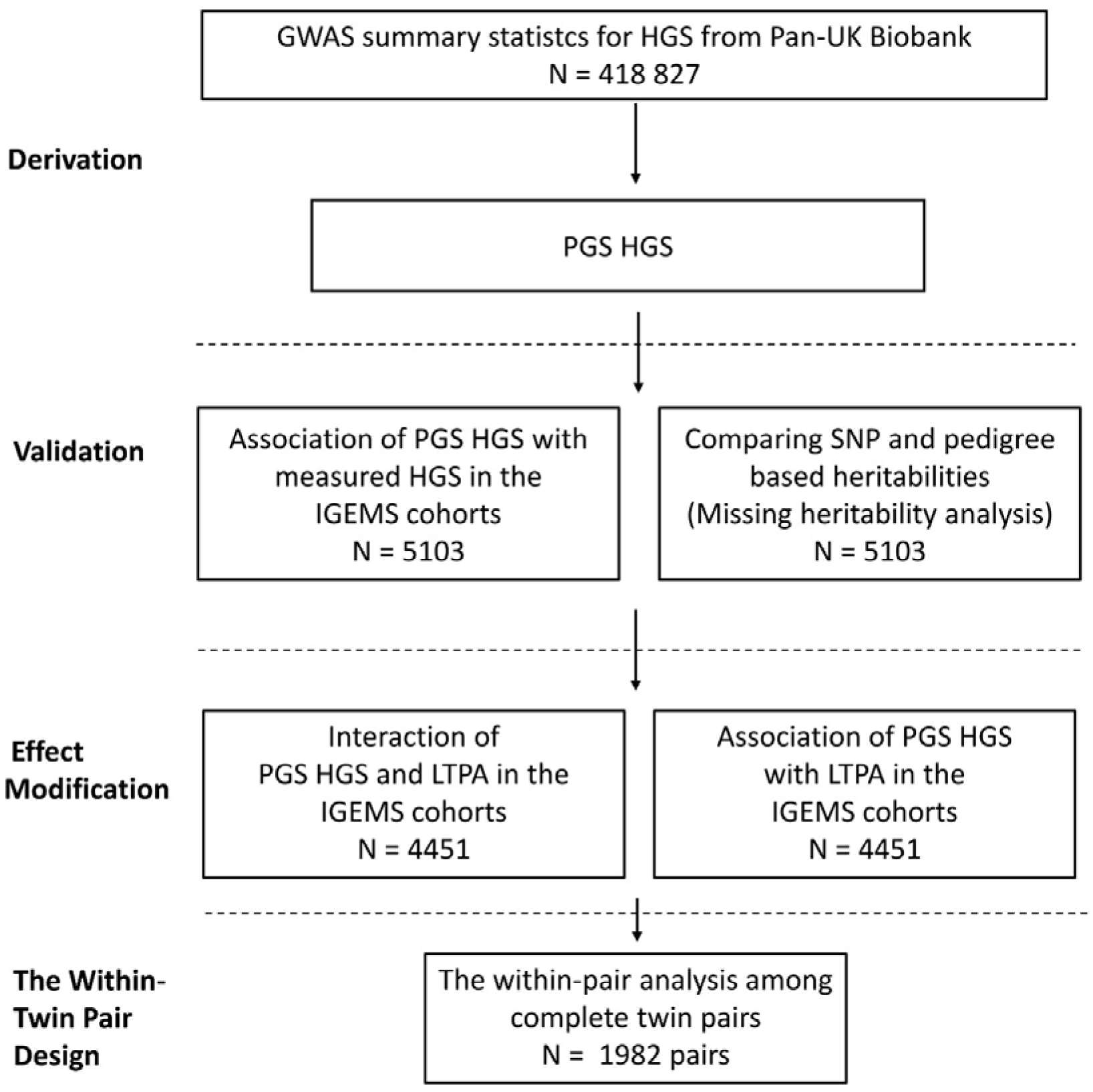
Study design and workflow. The polygenic score for handgrip strength (PGS HGS) was derived from the Pan-UK Biobank genome-wide association study summary statistics. Validation and effect modification analyses were conducted in the IGEMS cohorts, comprising a total of 5103 participants. A within-twin pair design was applied to 1982 complete dizygotic (DZ) and monozygotic (MZ) twin pairs. IGEMS, The Interplay of Genes and Environment across Multiple Studies; LTPA; leisure-time physical activity.

### Maximum HGS

In the UK Biobank (UKBB), maximal isometric HGS was measured using a calibrated hydraulic hand dynamometer (Jamar J00105; Lafayette Instrument Company, Lafayette, IN) (29). In all IGEMS studies, HGS was assessed by trained interviewers. Details of HGS measurement protocols in each IGEMS study are reported elsewhere (28,30–32). Due to the differing procedures for measuring HGS among the IGEMS studies, standardized maximum HGS was harmonized following IGEMS guidelines (28). Briefly, the sex- and study-specific standardized values for each wave were computed by normalizing the highest value obtained from all attempts within each wave based on the means and standard deviations (SDs) of maximum HGS from the first available waves in the respective studies. Given the recognized differences in HGS between men and women (11,12), we applied sex-specific standardized HGS values in our analysis. In the validation analysis, we used the first available HGS value for each participant. In the effect modification analysis, which included LTPA, we selected the HGS value closest in time to the LTPA measurement, prioritizing baseline assessments when available.

### Leisure-Time Physical Activity

LTPA includes all physical activities undertaken during free time, such as exercise, walking, heavy housework, and gardening (33). In the IGEMS studies, LTPA was self-reported using various items, which differed substantially across cohorts. To harmonize the data, continuous T-scores (mean = 50, SD = 10) were calculated, with higher scores indicating greater physical activity (34). For this study, the first available LTPA score was used for each cohort; however, LTPA was not assessed in the OCTO-Twin cohort.

### Covariates

Participant age was defined as age at the time of HGS measurement. BMI was calculated as weight divided by height squared (kg/m²). Height and weight data were collected at each study wave. For this study, the BMI value closest to the HGS assessment was selected. Measured BMI values were prioritized; if unavailable, self-reported values were used. The first 10 genetic PCs were used as covariates to mitigate the effects of population stratification (27). Country of residence was also included as a covariate to account for differences in population characteristics and data collection procedures across the twin cohorts.

### Genotype Data

Details of the genotyping procedures are provided in the Supplementary Materials under *Genotyping, quality control, and imputation*.

### Polygenic Score for HGS

PGS HGS was calculated for the IGEMS cohorts as a weighted sum of risk alleles, using effect sizes derived from GWAS summary statistics and the pipeline described in our previous study (24). The LD reference panel, summary statistics, and target sample were restricted to 1 006 473 common variants (MAF >5%) from the HapMap3 (35) reference panel, excluding the MHC region on chromosome 6 (6p22.1–21.3) in FITSA. These variants are typically well imputed in populations of European and Finnish ancestry. In the remaining IGEMS cohorts, we selected HapMap3 (35) SNPs with good imputation quality (INFO > 0.8) and MAF >1% across all cohorts, resulting in 952 885 SNPs. Using these SNP sets, the summary statistics were processed with the Bayesian method SBayesR (36), employing a linkage disequilibrium (LD) reference panel derived from a random sample of 50 000 UKBB (37) individuals to weight the GWAS effects. The final PGS was computed separately in each IGEMS cohort using these SBayesR-derived weights. The distribution of PGS HGS across the IGEMS cohorts is shown in Supplementary eFigure 3 and eFigure 4.

## Statistical Analyses

### Validation

#### Missing Heritability Analysis

Missing heritability, referring to the heritable factors not captured by SNPs in PGS HGS, was analyzed using the GCTA-GREML method, following the approach of Zaitlen et al. (38) and Yang et al. (39). Heritabilities were calculated with two genetic relatedness matrices: one accounting for family structure and the other for SNP genotypes. The difference between these estimates represents missing (or hidden) heritability. We estimated these components separately within each country. However, due to relatively small sample sizes in individual cohorts, the resulting SNP-based heritability estimates showed limited reliability (results not shown). Although the individual cohort estimates were not statistically significant (*p*>0.05), the pedigree-based heritability estimates were broadly consistent in magnitude with those reported in previous studies (13). Therefore, we proceeded with a meta-analysis of these estimates to improve the stability and interpretability of the findings across cohorts, using an inverse-variance weighted random-effects model.

#### Association Between PGS HGS and HGS

First, individuals were divided into PGS deciles to illustrate the linear association between standardized (mean = 0, SD = 1) PGS HGS and sex-specific standardized HGS. This association was analyzed using linear regression. Second, the proportion of variation in HGS explained by PGS HGS was examined using linear mixed models. Model predictors included age, country, the first 10 genetic PCs, and PGS HGS. Additionally, within-pair dependency was accounted for by including the family identifier as a random effect in all individual-level mixed models. Since the association between BMI and HGS is inconsistent across populations (40), BMI was not included as a covariate in the primary association analysis. However, as a sensitivity analysis, we repeated the primary association analysis between PGS HGS and HGS with additional adjustment for BMI, to assess whether the inclusion of BMI as a covariate influenced the observed associations. Results are reported as the total variance explained by the model (R²) and the change in R² (ΔR²) when PGS HGS was added to the model after the other predictors. Analyses were conducted for the full sample and separately by sex. Additionally, we fitted two interaction models to test whether the association between PGS HGS and HGS varied by age and country of residence. These models included PGS HGS × age and PGS HGS × country interaction terms, respectively, along with the main effects of age, country, and the first 10 genetic PCs. To evaluate country-specific differences in genetic associations, the PGS HGS × country interaction analysis was repeated with rotating reference groups, allowing for estimation of marginal effects within each country. Given evidence of heterogeneity across countries, we also fitted separate models within each country to examine country-specific associations between PGS HGS and HGS.

### Effect Modification by LTPA

#### Interaction Between PGS HGS and LTPA in HGS

To investigate whether LTPA may act as a moderator of the association between PGS HGS and HGS, we extended the association model to include both the main effect of LTPA and an interaction term PGS HGS × LTPA, representing a gene–environment interaction (41). This interaction term tested whether the effect of genetic predisposition on HGS varies depending on the level of LTPA, or conversely, whether the effect of LTPA on HGS differs across levels of genetic predisposition. BMI was included in this effect modification analysis, given its known association with reduced PA (42) and its possible biological relevance in modulating the genetic expression of muscular strength.

#### Mediation by LTPA in the PGS HGS-HGS Association

To evaluate LTPA as a potential mediator in the PGS HGS–HGS relationship, we also analyzed the association between PGS HGS and LTPA using linear mixed models adjusted for age at LTPA assessment, sex, country, and the first 10 genetic PCs. As PGS HGS was not statistically significantly associated with LTPA (β = 0.141, SE = 0.166, *p* = 0.392), further mediation analysis was not warranted.

### Within-twin-pair analysis

To further investigate the role of environmental factors, we conducted a fixed-effect within-twin-pair regression analysis (25) for all pairs and separately for DZ and MZ pairs. Finding an association between LTPA and HGS, particularly within MZ twins, supports an independent effect of LTPA, beyond shared genetic and environmental influences. Additionally, to explore whether genetic predisposition modifies the relationship between LTPA and HGS, we included a PGS HGS × LTPA interaction term in the model for DZ twins. For illustrative purposes, we examined marginal effects by categorizing PGS HGS into low, intermediate, and high levels of genetic liability, based on its tertile distribution. Because MZ twins are genetically identical and lack within-pair variation in PGS, an interaction term between PGS and LTPA was not estimated in this group. Instead, we plotted the regression coefficients for the association between LTPA and HGS, stratified by PGS HGS tertiles, among MZ twins to illustrate potential differences in effect size across levels of genetic liability. All fixed-effect within-twin-pair models were adjusted for age and BMI to account for confounding and any residual age differences due to non-simultaneous assessments.

Analyses were conducted using IMB SPSS Statistics, version 30.0.0 (172), R, version 4.2.3, and Stata, version 18.0, software. PGSs and PCs were calculated using Plink 2.0. Statistical significance was set at *p* < 0.05.

## Results

### Validation

Demographic characteristics of participants from the IGEMS studies included in the validation analysis are presented by country and sample in Table 1. The mean age of participants was 62.9 years (SD = 10.3), and of the 5103 participants, 44% were females.

**TABLE 1.**
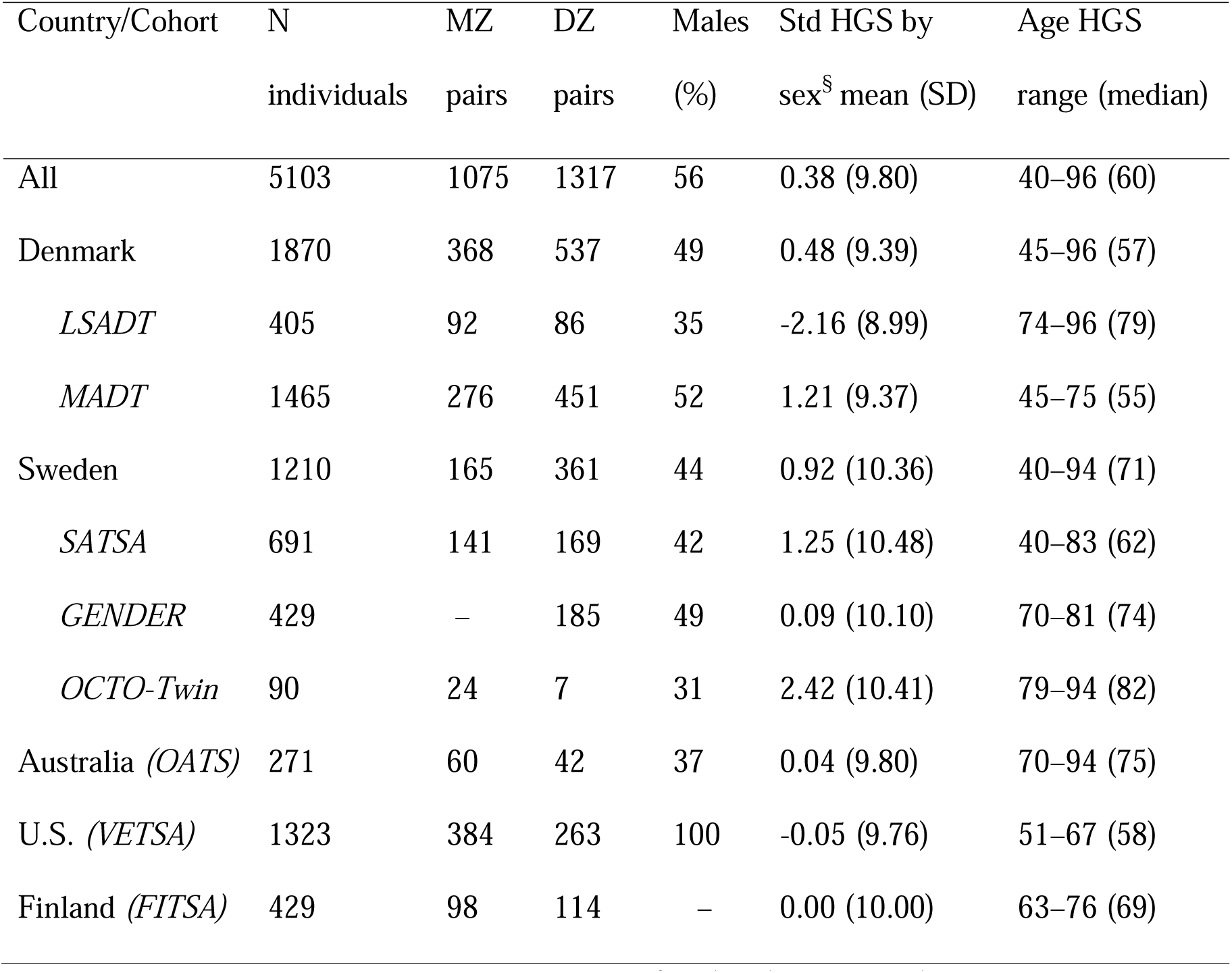
Sample characteristics by study at the first handgrip strength assessment. Note:§ the first possible handgrip measurement and corresponding age. MZ, monozygotic; DZ, dizygotic; HGS, handgrip strength; SE, standard deviation; U.S., United States.

#### Missing Heritability Analysis

Supplementary eFigure 5 shows the meta-analysis of pedigree-based heritability estimates across the IGEMS cohorts. The pooled results indicate a statistically significant difference between heritabilities (I² = 95%, *p* < 0.001), heritability estimates ranging from the lowest observed in Sweden (16%) and the highest in Australia (80%), while Finland, Denmark and the United States showed intermediate values (42%, 61% and 70%, respectively).

#### Association Between PGS HGS and HGS

The measured HGS increased linearly across deciles of PGS HGS in the IGEMS cohorts (R^2^ = 0.092, β = 0.714, SE = 0.052, *p* < 0.001) (Figure 2). Overall, PGS HGS accounted for 4.6% of the variation (ΔR^2^) in measured HGS (Table 2). When stratified by sex, the explained variance was 4.3% in males and 5.2% in females. Sensitivity analyses, including BMI as a covariate, were consistent with our primary results (Supplementary eTable 1 and eTable 2). No statistically significant interaction between PGS HGS and age was observed in the overall sample (β = –0.016, SE = 0.014, *p_interaction_*= 0.258), nor in males (β = -0.006, SE = 0.021, *p_interaction_* = 0.779) or females (β = -0.038, SE = 0.020, *p_interaction_*= 0.058; Supplementary eTable 3). However, a statistically significant interaction between PGS HGS and country of residence was detected in the PGS HGS × country model, using Australia as the reference group (Supplementary eTable 4). Specifically, Denmark (*p_interaction_* = 0.031), Sweden (*p_interaction_* = 0.009), and the United States (*p_interaction_* = 0.009) showed interaction effects, suggesting that the strength of the association between PGS HGS and measured HGS differs across countries (Supplementary eFigure 6). Finland did not differ significantly (*p_interaction_* = 0.174). No statistically significant interactions were observed when other countries were used as the reference group, reinforcing that the primary contrasts were with Australia. In country-stratified models, PGS HGS explained between 2.9% and 7.7% of the variation in measured HGS, with the weakest association in Sweden and the strongest in Australia (Table 2). When further stratified by sex, the variance explained ranged from 2.0% (Swedish males) to 6.3% (Australian males), and from 3.8% (Swedish females) to 8.6% (Australian females).

**Figure 2.**
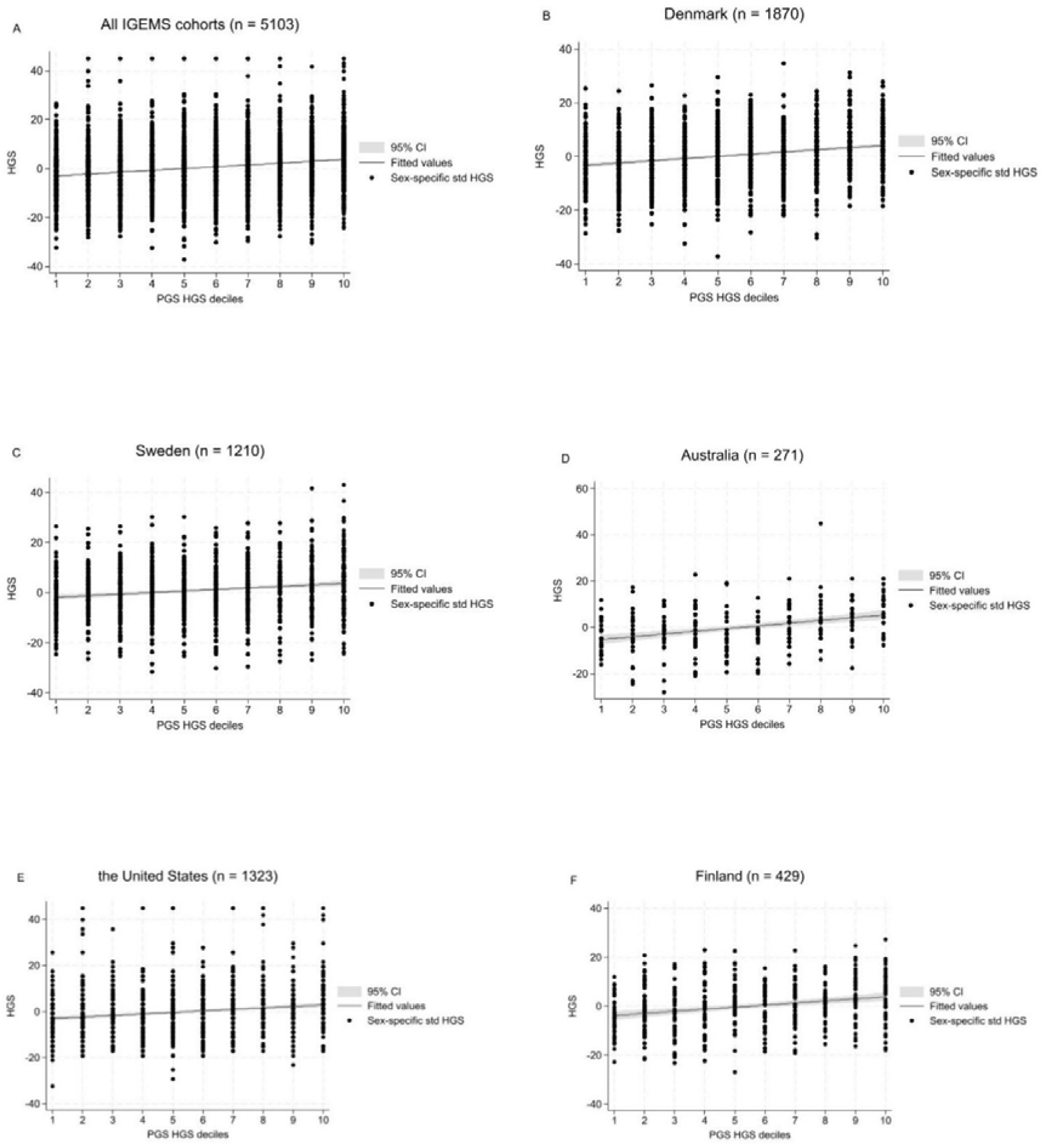
Association between a PGS HGS deciles and measured HGS. Fitted values from a linear regression of HGS on PGS HGS deciles. Panel A shows the association across all IGEMS cohorts, Panel B in the Danish cohorts, Panel C in the Swedish cohorts, Panel D in the Australian cohort, Panel E in the United States cohort, and Panel F in the Finnish cohort. PGS, polygenic score; HGS, handgrip strength; CI, confidence interval.

**TABLE 2.** Associations between a polygenic score for handgrip strength (PGS HGS) and isometric handgrip strength in the IGEMS cohorts.

|  | zPGS HGS |  |  | Full model |  |  |  |
| --- | --- | --- | --- | --- | --- | --- | --- |
| | beta | SE | *P* | $R^2$ (%) | *P* | $\Delta R^2$ (%) | N |
| All | 2.14 | 0.15 | <0.001 | 9.6 | <0.001 | 4.6 | 5103 |
| Males | 2.05 | 0.19 | <0.001 | 10.2 | <0.001 | 4.3 | 2858 |
| Women | 2.33 | 0.23 | <0.001 | 10.3 | <0.001 | 5.2 | 2245 |
| Denmark | 2.25 | 0.23 | <0.001 | 15.3 | <0.001 | 5.6 | 1870 |
| Males | 2.25 | 0.32 | <0.001 | 19.2 | <0.001 | 5.9 | 907 |
| Women | 2.23 | 0.33 | <0.001 | 14.1 | <0.001 | 5.3 | 963 |
| Sweden | 1.76 | 0.32 | <0.001 | 6.4 | <0.001 | 2.9 | 1210 |
| Males | 1.44 | 0.46 | <0.001 | 6.8 | <0.001 | 2.0 | 528 |
| Women | 2.08 | 0.42 | <0.001 | 7.8 | <0.001 | 3.8 | 682 |
| Australia | 2.84 | 0.50 | <0.001 | 34.8 | <0.001 | 7.7 | 271 |
| Males | 2.65 | 0.83 | 0.002 | 56.5 | <0.001 | 6.3 | 100 |
| Women | 2.97 | 0.67 | <0.001 | 26.9 | <0.001 | 8.6 | 171 |
| United States (Males) | 1.98 | 0.29 | <0.001 | 5.8 | <0.001 | 4.0 | 1323 |
| Finland (Females) | 2.48 | 0.54 | <0.001 | 9.39 | 0.002 | 6.1 | 429 |

Note: The results are shown for the full model including zPGS HGS as a predictor and adjusted for age and 10 principal genetic components. Analysis in full cohort was additionally adjusted for country. HGS was standardized by sex. Family number was
included in the models as a random factor. $\Delta R^2$ describes difference in coefficient of determination ( $R^2$ ) between adjusted model with and without PGS HGS. z, standardized for normal distribution; PGS, polygenic score; HGS, handgrip strength; IGEMS, The Interplay of Genes and Environment across Multiple Studies; SE, standard error.

### Effect Modification by LTPA

The demographic characteristics of participants included in the effect modification analysis are presented by country and sample in the Supplemental eTable 5. The distributions of LTPA are shown in Supplementary eFigure 7 and eFigure 8. A statistically significant interaction between PGS HGS and LTPA was observed in the full sample (β = -0.034, SE = 0.014, *p_interaction_* = 0.013), in females (β = -0.043, SE = 0.020, p_interaction_ = 0.035), as well as in the Australian cohort (total sample: β = -0.182, SE = 0.065, *p_interaction_* = 0.006; men: β = -0.306, SE = 0.119, *p_interaction_* = 0.013; women: β = -0.240, SE = 0.081, *p_interaction_* = 0.004; Supplementary eTable 6). These results suggest that the positive effect of LTPA on HGS was more pronounced among individuals with a genetic predisposition for lower muscle strength (Figure 3 and Supplementary eFigure 9).

**Figure 3.**
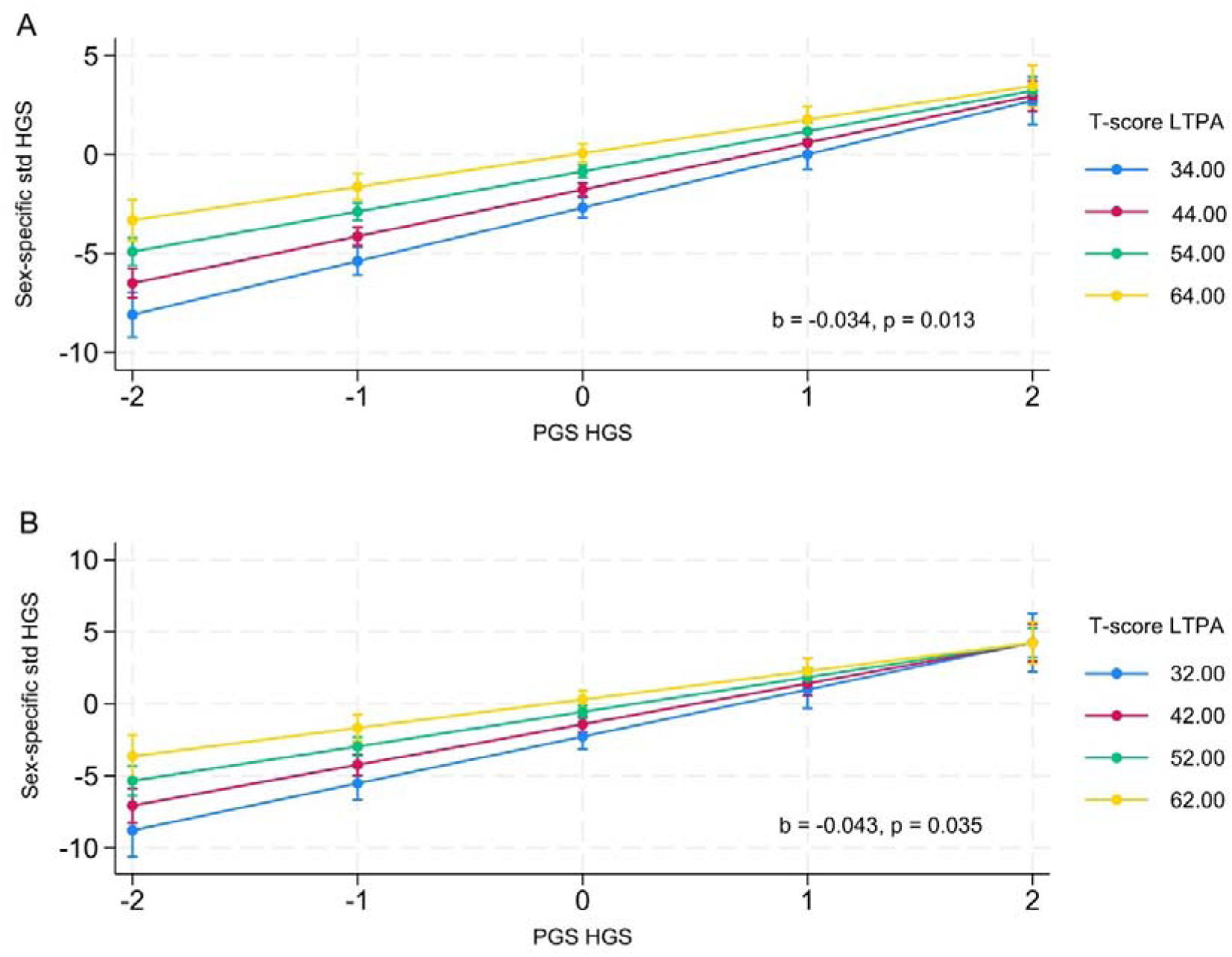
Interaction between polygenic score for handgrip strength (PGS HGS) and leisure-time physical activity (LTPA) on sex-specific standardized handgrip strength across all cohorts. The figure shows that the positive effect of LTPA on HGS was stronger among individuals with a lower PGS HGS. The results are based on linear regression models incorporating the main effects of PGS HGS and LTPA, along with their interaction term (PGS HGS × LTPA). Models are adjusted for age at HGS measurement, BMI, country of residence, and 10 principal genetic components, with family number included as a random factor. Both PGS HGS and LTPA were treated as continuous variables in the models. For illustrative purposes, the figure depicts T-score cut-off points, where the lowest value represents the 5th percentile, the highest value represents the 95th percentile, and intermediate values are spaced at 10-point intervals. Panels represent: (A) all cohorts combined; (B) females in all cohorts. Beta coefficients (b) and p-values indicate the strength and statistical significance of the PGS HGS × LTPA interaction term. The plotted lines represent estimated associations, and the error bars indicate 95% confidence intervals (CIs).

### Within-twin-pair analysis

Descriptive characteristics of the twin pairs included in the within-pair analysis are presented in Supplementary eTable 7, with means and standard deviations of within-pair differences shown in Supplementary eTable 8. Results from the fixed-effects within-twin-pair regression analysis are detailed in Supplementary eTable 9. Higher levels of LTPA were associated with greater HGS in the full sample of twin pairs (β = 0.059, *p* = 0.001) and among DZ twins (β = 0.090, *p* < 0.001), but attenuated among MZ twins (β = 0.004, *p* = 0.894). Although the interaction between PGS HGS and LTPA was not statistically significant in DZ twins (β = – 0.008, *p_interaction_* = 0.752), both the marginal effects plot for DZ twins (Supplementary eFigure 10) and the stratified regression coefficients for MZ twins (Supplementary eFigure 11) suggest a weak but consistent trend indicating that individuals with a genetic predisposition for lower HGS may benefit more from higher LTPA than those genetically predisposed to greater muscle strength.

## Discussion

In this multinational twin study, we aimed to extend the evaluation of a PGS HGS to diverse international cohorts and to investigate whether its association with measured strength is modified by age, sex, country of residence, or LTPA. Our findings confirmed that higher PGS HGS was associated with greater measured HGS, with modest differences across countries. The association remained consistent in sex-stratified models, accounting for slightly more variance in females than in males. Notably, we observed a statistically significant interaction between PGS HGS and LTPA, suggesting that individuals with a lower genetic predisposition for higher muscular strength may benefit more from LTPA. While our within-twin pair analyses offered limited support for an independent environmental effect of LTPA, the observed gene–environment interaction underscores the potential of PA to attenuate genetic disadvantage in muscle strength.

Our findings align with existing literature, suggesting that HGS is moderately influenced by genetic factors, with heritability estimates (h^2^) ranging from 30% to 65% in classical twin studies (13). However, our meta-analysis of pedigree-based heritability estimates across the IGEMS cohorts revealed substantial variability, with estimates ranging from 16% in Sweden to 80% in Australia. The Australian estimate appears relatively high, whereas previous twin studies have reported heritability estimates for HGS in the Swedish cohort ranging from 47% in females to 72% in males (11). Classic twin-based designs may overestimate heritability due to factors such as genetic interactions, including epistasis and dominance, gene– environment interactions, or violations of the equal environments assumption (38,43). Conversely, pedigree-based GCTA-GREML methods typically yield more conservative estimates (38). Yet our dataset included only MZ and DZ twin pairs, limiting the range of genetic relatedness. This constraint may reduce the method’s ability to distinguish additive genetic effects from non-additive genetic and environmental influences, potentially inflating the heritability estimates. Additionally, the relatively small sample sizes, particularly in the Australian cohort, resulted in low statistical power, likely contributing to inflated point estimates and underscoring the need for caution in interpretation.

While heritability estimates quantify the proportion of trait variance attributable to genetic differences in a certain population, the predictive power of PGS reflects how much of that variance can be captured using the common SNPs identified in GWASs, under the assumption of additive genetic effects only (44). In the present study, the PGS HGS was significantly associated with measured strength, explaining 4.6% of the variance overall. While this proportion reflects only a fraction of the total heritable variation, it is consistent with the modest predictive power typically observed for PGSs of complex traits (20,44). Extending our previous findings in Finnish females, where the PGS HGS explained 6.1% of the variation in measured strength (24), the present results demonstrate similar predictive utility across international cohorts, with variance explained ranging from 2.9% to 7.7% in country-stratified models. The strongest associations were observed in Australia, particularly among females (8.6%), while the weakest were in Sweden. These findings reinforce the generalizability of the PGS but may also reflect differences in cohort composition, environmental exposures, or gene–environment correlations across countries (20,45). Notably, the higher predictive power observed in the Australian cohort may be partly attributable to shared genetic ancestry with the discovery sample used to build the PGS, as the original GWAS was based on the UK Biobank, and a substantial proportion of the Australian population has British or Irish ancestry (46). Conversely, the lower estimates in the Swedish cohorts may reflect more selective sampling. For example, the OCTO-Twin study included only like-sexed twin pairs aged 80 years or older who were both still alive at recruitment (47), and the GENDER study comprised older, unlike-sex twin pairs (48), introducing potential biases due to age, survival, and cohort effects.

Genetic contributions to HGS have generally been found to be similar across sexes (12,13), though some prior research has pointed to a stronger heritable component in males and a greater influence of shared environmental factors in females (11,16). Additionally, one study found that a genetic score based on 16 HGS-associated SNPs predicted the phenotype more effectively in men than in women (19). In contrast, we observed slightly higher variance explained by the PGS HGS in females (5.2%) than in males (4.3%), even if the overall performance was broadly comparable between sexes across all cohorts. The extent to which genetic effects on HGS are stable across adulthood also remains debated (11–15). While some evidence suggests age-related changes in genetic effects (13,15,23), we found no indication that the predictive validity of the PGS diminished with increasing age. This implies that the genetic architecture underlying HGS remains relatively stable across the adult lifespan, despite theoretical expectations that accumulating environmental exposures may weaken genetic influences over time (49). Sex-specific exposures, hormonal differences, and differential life expectancy may introduce additional environmental variability, particularly in women, which could obscure genetic effects (17,31). However, we found no strong evidence for meaningful sex differences in the predictive power of the PGS. Rather than reflecting underlying differences in genetic architecture, the modest sex variation observed in our study may result from cohort-specific characteristics, the specific variants included in the PGS, or limited power in sex-stratified analyses. Future research, including sex-stratified genome-wide association studies, may provide deeper insights into the mechanisms underlying sex differences in the genetic basis of HGS (22).

While robust detection of gene–environment interactions is generally difficult (41,50), our findings provide evidence for a gene–environment interaction between LTPA and genetic predisposition for HGS. Specifically, the positive association between LTPA and HGS was more pronounced among individuals with a lower PGS HGS. This pattern aligns with the differential susceptibility framework, which posits that individuals with certain genetic profiles may be more sensitive to environmental influences, and thus more responsive to lifestyle interventions like PA (51). Our findings also support the previous research suggesting that PA can partially compensate for the genetic risk in various health-related traits (52,53). However, results from our within-twin pair analysis did not support a causal role of PA in promoting muscle strength. While LTPA was associated with greater HGS within DZ twin pairs, this association was not observed among MZ twins. The attenuation of the LTPA– HGS relationship in MZ twins implies that the association is at least partly attributable to shared genetic influences. Nonetheless, the observed trends in stratified and marginal effects, though not statistically significant, indicated that even among genetically identical individuals, higher levels of activity may offer some benefit, particularly for those with lower genetic predisposition. Furthermore, while LTPA moderated the relationship between genetic predisposition and HGS, it did not mediate it. The lack of a significant association between PGS HGS and LTPA supports our previous notion that genetic predisposition for strength does not substantially influence individuals’ engagement in LTPA (54). However, it is important to note that PA itself is a heritable trait, with genetic factors accounting for a moderate proportion of individual differences in PA behavior (55). This raises the possibility that the association between LTPA and HGS may be confounded by shared genetic influences affecting both traits. Such pleiotropic effects could explain why the association between LTPA and HGS was attenuated in MZ twins, who are genetically identical and typically show high concordance in PA behavior, thereby limiting detectable variation in exposure. Taken together, our results underscore the complex interplay between genetic and environmental influences on muscular strength. Future studies should apply integrative analytical approaches that account not only for the main effects of genetic variants but also for their potential interactions with environmental exposures (50), such as PA, to better elucidate individual variability in response to lifestyle interventions.

This study has several strengths and limitations. A key strength is the relatively large overall sample size (N > 5000) drawn from multinational cohorts, which enhances statistical power and enables replication across samples. HGS and LTPA were harmonized across cohorts using validated methods (28,34), allowing for comparability; however, this harmonization may have led to some loss of cohort-specific detail. LTPA was assessed via self-report questionnaires, which may be subject to recall or reporting bias. In addition, while the use of PGS HGS derived from the UK Biobank offers a robust genetic proxy, it may limit the generalizability of findings to more diverse populations due to ancestry-related differences (20). Although the overall sample was sizable, individual cohort sizes were relatively small, potentially limiting power in stratified or interaction analyses. Furthermore, the cross-sectional nature of the primary analyses constrains causal inference, though the within-twin pair design partially mitigates this by accounting for shared genetic and environmental confounding (25). Finally, while PGS capture aggregate genetic risk, they do not reflect the full complexity of gene–environment interplay (50) and may miss rare or structural variants contributing to muscular strength (20).

## Conclusion

Our findings confirm that PGS HGS holds value for identifying population-level genetic influences on muscle strength. Importantly, while genetic factors set a baseline, lifestyle behaviors such as PA may partially offset lower genetic predisposition in terms of muscle strength, supporting the role of PA as a modifiable factor in maintaining physical fitness. These results underscore the importance of promoting PA broadly, and they point to the potential utility of integrating genetic information in tailoring future health interventions.

## Acknowledgments

We thank the IGEMS and UK Biobank study participants for their valuable contribution to science, as well as the researchers involved in the original data collection. We also thank data managers Patricia St. Clair and Orla Hayden from the University of Southern California for their essential support with data access and processing. We are grateful to research assistant Joseph White from the University of Colorado Boulder for his valuable assistance with data analysis.

## Funding

This study was funded by the Research Council of Finland (grant numbers: 341750, 346509, and 361981 to E.S., 336823 to J.K.), the Juho Vainio Foundation (E.S.), and the Päivikki and Sakari Sohlberg Foundation (E.S). Funding for the IGEMS studies is detailed in the Supplementary Materials under *Funding for the IGEMS Studies*.

## Conflict of Interests

None declared.

## Author Contributions

ES and PH conceived of the idea for the study, as well as contributed to the design of the study and interpreted the data. P.H., M.N., I.K.K., A.T., and J.W. performed the statistical analysis, assisted by T..P, C.A.R., J.K., and E.S. P.H. and E.S. drafted the first version of the manuscript. All authors reviewed the manuscript and revised it critically for important intellectual content. All of the authors have also approved the conducted analyses and the final version of the manuscript to be published. M.N., I.K.K., A.T., K.A.M., C.A.R., M.S.P., T.R., D.F., M.G., N.L.P., and J.K. contributed to IGEMS data collection and management. E.S. is a guarantor, and as the corresponding author, she attests that all listed authors meet authorship criteria and that no others meeting the criteria have been omitted.

## Data availability

Comprehensive information on data availability and access procedures is available in the Supplementary Materials under *Data availability for IGEMS studies*.

## Notes

### Competing Interest Statement

The authors have declared no competing interest.

### Author Declarations

The North West Multi-centre Research Ethics Committee approved the UK Biobank study (21/NW/0157). The IGEMS Consortium was approved by the University of Southern California Institutional Review Board (approval number UP-16-00315). The FITSA data collection was approved by the Ethics Committee of the Central Hospital District of Central Finland (KSSHP 24/2000). All studies obtained informed written consent from all participants and adhered to the principles of the Declaration of Helsinki.

### Summary of Updates

This version includes minor corrections to typographical errors in the manuscript. Supplemental files updated. No changes were made to the data, analyses, or conclusions.

