## Supplemental material for "Evaluating a Genome-Wide Polygenic Score for Handgrip Strength and Its Interplay with Leisure-Time Physical Activity Across the IGEMS Twin Cohorts"

eFigure 1. Participant Flow Chart of Individual Analysis

eFigure 2. Participant Flow Chart of Within-Twin Pair Analysis

Overview of IGEMS Cohorts Included in the Analysis

Genotyping, quality control, and imputation

eFigure 3. Density Curves of PGS HGS among the IGEMS Participants and Stratified by Sex

eFigure 4. Density Curves of PGS HGS among the IGEMS Participants Stratified by Country

eFigure 5. Forest Plot of the Meta-Analysis of Pedigree-Based Heritability Estimates across IGEMS Cohorts

eTable 1. Sample Characteristics by Study: Participants Included in the Sensitivity Analysis with Complete Case Data for the First Available HGS and Corresponding BMI Measurement (N = 5020)

eTable 2. Associations Between a PGS HGS and Isometric HGS among Participants with

Complete Case Data for the First Available HGS and Corresponding BMI Measurement (N = 5020)

eTable 3. Interactions Between zPGS HGS and Age

eTable 4. Interactions Between zPGS HGS and Country

eFigure 6. Marginal Effects of PGS HGS Within Countries from Interaction Models

eTable 5. Sample Characteristics of the Participants with Complete Case Data for HGS, BMI, and LTPA Measurements (N = 4451)

eFigure 7. Density Curves of LTPA among the IGEMS Participants and Stratified by Sex

eFigure 8. Density Curves of LTPA among the IGEMS Participants Stratified by Country

eTable 6. Interactions Between zPGS HGS and LTPA

eFigure 9. Interaction between PGS HGS and LTPA on sex-specific standardized HGS in Study II within the Australian cohort.

eTable 7. Descriptive Characteristics of the Twin Pairs Included in the Within-Twin Pair Analysis

eTable 8. Mean and Standard Deviation of Within-Pair Differences

eTable 9. The Association Between LTPA and HGS: Within-Twin Pair Analysis

eFigure 10. Marginal Effects of LTPA Within DZ Twin Pairs from Interaction Models

eFigure 11. The Stratified Regression Coefficients for MZ Twins by PGS HGS Tertiles

Funding for the IGEMS Studies

Data availability for IGEMS Studies

**eFigure 1. Participant Flow Chart of Individual Analysis**

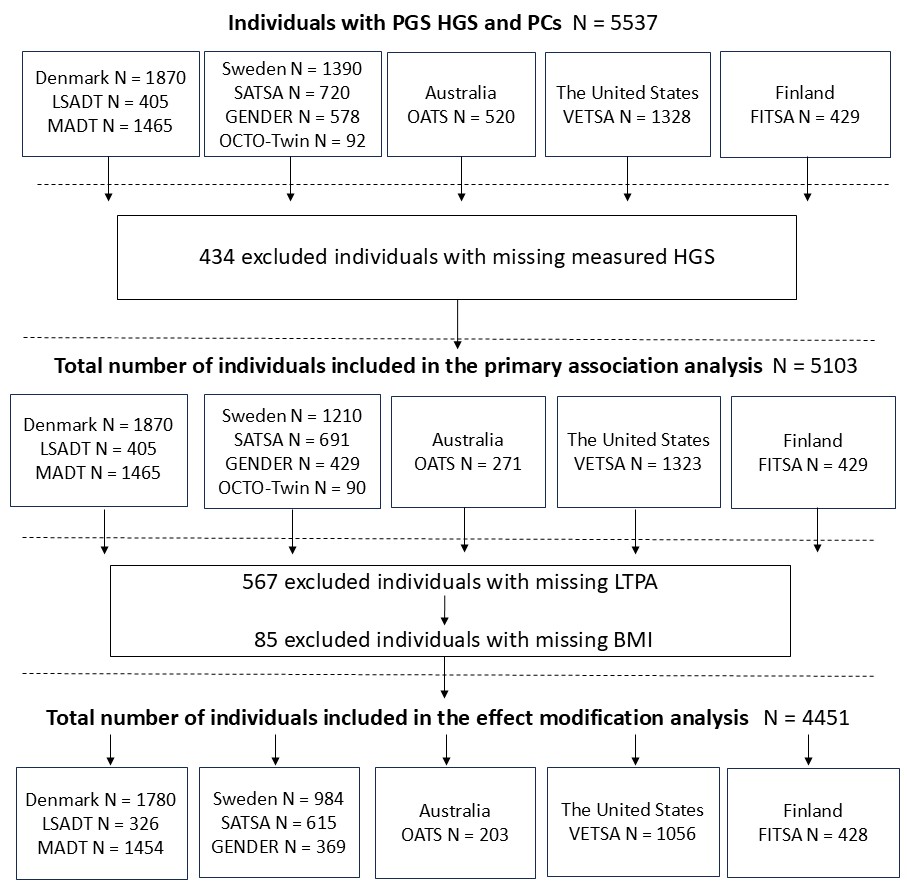

**eFigure 2. Participant Flow Chart of Within-Twin Pair Analysis**

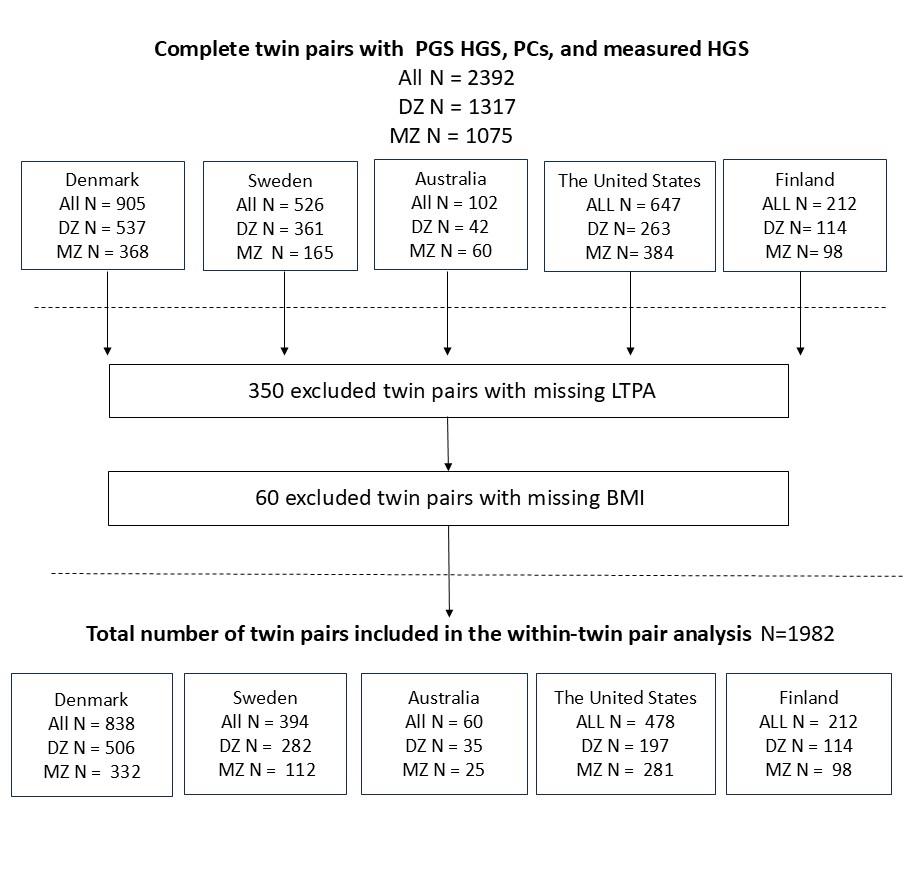

**Overview of IGEMS Cohorts Included in the Analysis**

*Denmark*

*The Longitudinal Study of Aging Danish Twins* (LSADT) (1) and *the study of Middle-Aged Danish Twins* (MADT) (2) were ascertained through the Danish Twin Registry and the Danish Central Person Registry, including both same- and opposite-sex twin individuals. LSADT was conducted over six waves from 1995 to 2005, all participants were born in Denmark before 1920. MADT was a sample of participants aged 46 to 68 years at baseline in 1998. Follow-up was performed in 2008-2011. We included 405 LSADT and 1465 MADT participants in our study.

*Sweden*

*The Swedish Adoption/Twin Study of Aging* (SATSA) (3), *Aging in Women and Men: A Longitudinal Study of Gender Differences in Health Behaviour and Health among Elderly* (GENDER) (4) , and *Origins of Variance in the Oldest Old: Octogenarian Twins* (The OCTO-Twin Study) (5) were derived from data originating in the population-based Swedish Twin Registry. SATSA was a longitudinal study with over 28 years of follow-up consisting of twins reared apart and a control sample reared together. The baseline questionnaire was conducted in 1984, and by the end of 2014, nine survey waves and 10 personal testing waves were finalized. GENDER began in 1995 with a survey for opposite-sex twin pairs, born between 1906 and 1925, with two follow-ups in 1999-2001 and 2003-2005. A final survey was conducted in 2008. The OCTO-Twin Study comprised like-sexed twin pairs who had surpassed the age of 80 at the starting point in 1991, and at the end of the study in 2002, five waves of data collection at 2-year intervals were completed. We examined a sample of 691 twin individuals from SATSA, 429 from GENDER, and 90 from the OCTO-Twin Study.

*Australia*

The Older Australian Twins Study (OATS) (6) included same- and opposite-sex twins aged 65 years or older at baseline 2007–2008 in the three Eastern states of Australia. In-person assessments follow participants every two years. The second follow-up was accomplished between 2009 and 2013, the third was between 2012 and 2016. Our study sample included 271 twins from OATS.

*The Unites States*

*The Vietnam Era Twin Study of Aging* (VETSA) (7) included male twin pairs who served in the US Military between 1965 and 1975. Participants were randomly selected from the larger Vietnam Era Twin Registry and were in their 50s at the time of the recruitment. The first wave began in 2003. Follow-ups are conducted every five years, and the third wave was completed in 2019. A total of 1323 twins from VETSA, including attrition-replacement participants who were assessed for the first time in the second wave, were part of our study sample.

*Finland*

*The Finnish Twin Study of Aging* (FITSA) (8) is a longitudinal study drawn from the older Finnish Twin Cohort (9). It included MZ and DZ female twin pairs aged 63–76 years at baseline in 2000–2001. Follow-ups were conducted in 2003–2004 (in-person and postal) and later via postal questionnaire. We included 429 participants from the baseline wave in our study.

**Genotyping, quality control, and imputation**

In the UKBB study, genome-wide genotyping was performed using the UKBB Axiom Array, which includes coding variants across a range of minor allele frequencies (MAF), including both rare variants (<1% MAF) and markers that ensure good genome-wide coverage for imputation in European populations within the common (>5%) and low-frequency (1%–5%) MAF ranges. Detailed information on genotyping, quality control, and imputation in the UKBB is provided in the UKBB documentation (10,11). In the Finnish cohort (the older FTC, including FITSA), genotyping was conducted using the Illumina Human610-Quad v1.0 B, Human670-QuadCustom v1.0 A, several versions of the Illumina HumanCoreExome array (12 v1.0 A, 12 v1.1 A, 24 v1.0 A, 24 v1.1 A, 24 v1.2 A), and the Affymetrix FinnGen Axiom array (12). Variants with MAF <1% and Hardy–Weinberg equilibrium P-values <1 × 10⁻⁶ were excluded. Imputation was performed using the Haplotype Reference Consortium (HRC) reference panel (release 1.1) (13). In the remaining IGEMS cohorts, genotyping was conducted within each study using established procedures, primarily with the Illumina OmniExpress and Infinium PsychArray BeadChip platforms. Other arrays used include the Illumina 670k and HumanCoreExome. All data were imputed to the 1000 Genomes Project reference panel (Phase 1, Version 3 or Phase 3, Version 5) (14) and/or the Haplotype Reference Consortium panel (13).

**eFigure 3. Density Curves of PGS HGS among the IGEMS Participants and Stratified by Sex**

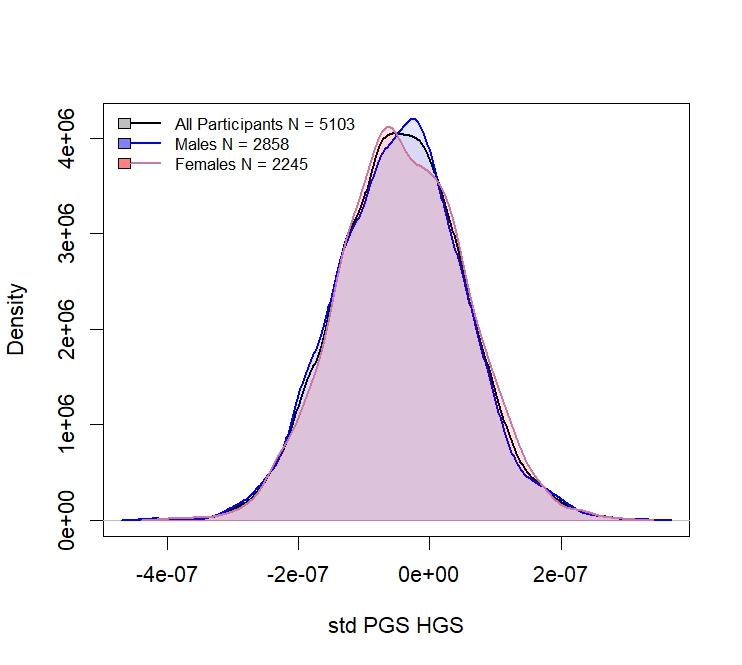

**eFigure 4. Density Curves of PGS HGS among the IGEMS Participants Stratified by Country**

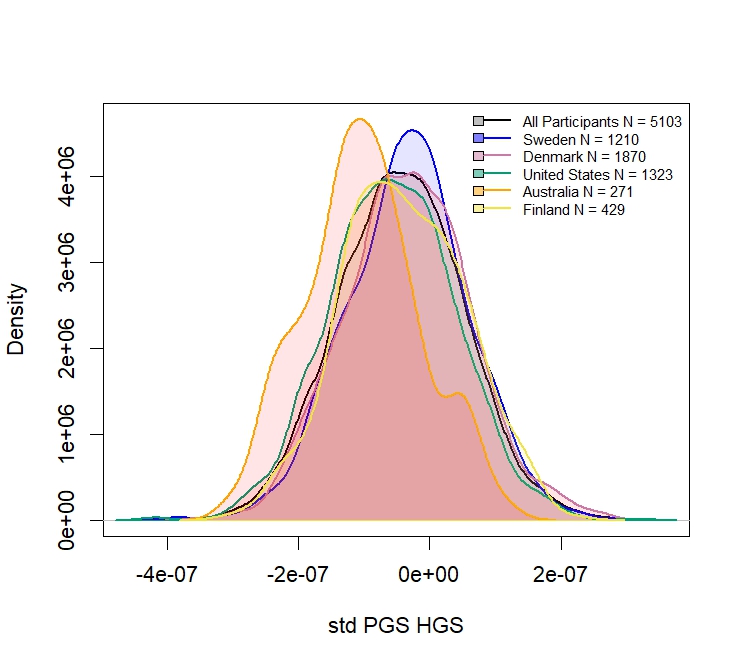

**eFigure 5. Forest Plot of the Meta-Analysis of Pedigree-Based Heritability Estimates across IGEMS Cohorts**

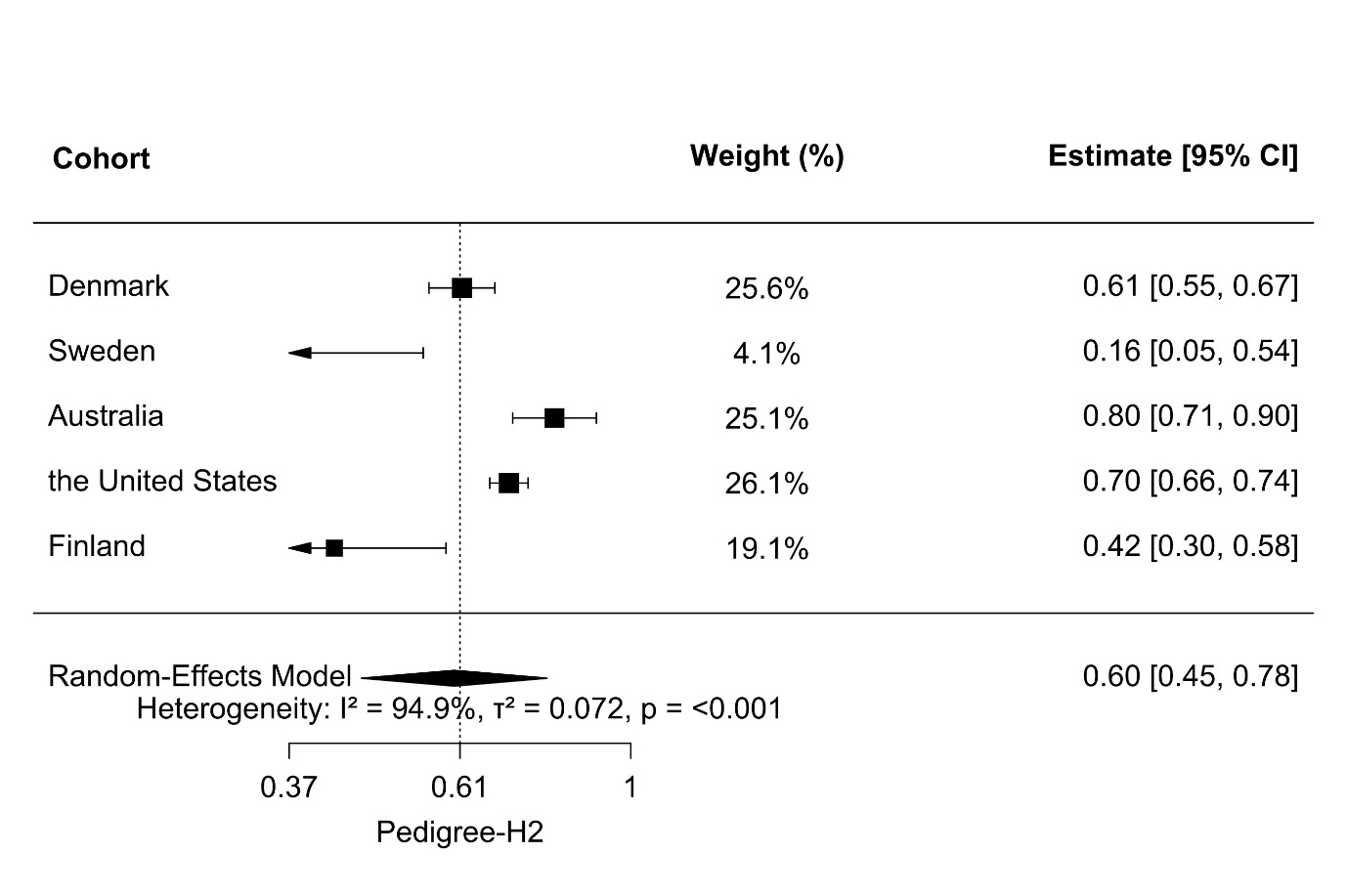

The effect sizes represent variance estimates back-transformed from the logarithmic scale (exponentiated values). The squares indicate cohort-specific estimates, with their sizes reflecting the weight of each study in the meta-analysis. Horizontal lines show 95% confidence intervals (CIs). The diamond represents the overall pooled estimate from the random-effects model, with its width indicating the corresponding 95% CI. Heterogeneity statistics (I², τ², and p-value) are reported below the plot.

**eTable 1. Sample Characteristics by Study: Participants Included in the Sensitivity Analysis with** **Complete Case Data for the First Available HGS and Corresponding BMI Measurement (N = 5020)**

| Country/Cohort | N | Std HGS by sex^§^  mean (SD) | Age HGS  range (median) | BMI^†^  mean (SD) | Age BMI  range (median) |
| --- | --- | --- | --- | --- | --- |
| All | 5020 | 0.46 (9.77) | 40–96 (60) | 26.77 (4.63) | 40–98 (60) |
| Denmark | 1791 | 0.71 (9.28) | 45–96 (57) | 24.96 (3.52) | 45–98 (57) |
| *LSADT* | 326 | -1.56 (8.52) | 74–96 (78) | 24.60 (3.67) | 76–98 (80) |
| *MADT* | 1465 | 1.21 (9.37) | 45–75 (55) | 25.04 (3.48) | 45–75 (55) |
| Sweden | 1208 | 0.93 (10.35) | 40–94 (71) | 25.96 (3.94) | 40–94 (71) |
| *SATSA* | 691 | 1.25 (10.48) | 40–83 (62) | 25.81 (3.91) | 40–83 (62) |
| *GENDER* | 427 | 0.12 (10.11) | 70–81 (74) | 26.52 (3.93) | 71–80 (74) |
| *OCTO-Twin* | 90 | 2.42 (10.41) | 80–94 (82) | 24.42 (3.74) | 56–94 (82) |
| Australia *(OATS)* | 271 | 0.04 (9.80) | 70–94 (75) | 27.14 (4.51) | 70–94 (75) |
| U.S. *(VETSA)* | 1322 | -0.05 (9.77) | 51–67 (58) | 29.52 (3.34) | 51–67 (58) |
| Finland (*FITSA*) | 428 | -0.02 (10.00) | 63–76 (69) | 27.93 (4.73) | 63–76 (69) |

Note: § the first available HGS measurement and corresponding age;†the BMI measurement and corresponding age closest to the HGS assessment; HGS, handgrip strength; BMI, body mass index; SD, standard deviation.

**eTable 2. Associations Between a PGS HGS and Isometric HGS among Participants with**

**Complete Case Data for the First Available HGS and Corresponding BMI Measurement (N = 5020)**

|  |  |  | Models without BMI as a covariate | | | | |  | Models with BMI as a covariate | | | | |  |
| --- | --- | --- | --- | --- | --- | --- | --- | --- | --- | --- | --- | --- | --- | --- |
|  |  |  | zPGS HGS | | | Full model | |  | zPGS HGS | | | Full model | |  |
|  |  | N | β | SE | *P* | R^2^ (%) | *P* | ΔR^2^ (%) | β | SE | *P* | R^2^ (%) | *P* | ΔR^2^ (%) |
| All | | 5020 | 2.16 | 0.15 | <0.001 | 9.1 | <0.001 | 4.7 | 2.17 | 0.15 | <0.001 | 9.5 | <0.001 | 4.7 |
| Males | | 2829 | 2.04 | 0.19 | <0.001 | 9.8 | <0.001 | 4.3 | 2.05 | 0.19 | <0.001 | 10.5 | <0.001 | 4.4 |
| Females | | 2191 | 2.38 | 0.23 | <0.001 | 10.0 | <0.001 | 5.4 | 2.39 | 0.23 | <0.001 | 10.1 | <0.001 | 5.5 |
| Denmark | | 1791 | 2.29 | 0.24 | <0.001 | 14.3 | <0.001 | 6.0 | 2.33 | 0.24 | <0.001 | 15.6 | <0.001 | 6.2 |
| Males | | 880 | 2.22 | 0.33 | <0.001 | 18.1 | <0.001 | 5.9 | 2.18 | 0.33 | <0.001 | 20.1 | <0.001 | 5.7 |
| Females | | 911 | 2.33 | 0.34 | <0.001 | 13.7 | <0.001 | 5.9 | 2.40 | 0.34 | <0.001 | 4.4 | <0.001 | 6.2 |
| Sweden | | 1208 | 1.75 | 0.32 | <0.001 | 6.3 | <0.001 | 2.8 | 1.76 | 0.32 | <0.001 | 6.3 | <0.001 | 2.8 |
| Males | | 527 | 1.41 | 0.46 | <0.001 | 6.8 | <0.001 | 1.9 | 1.42 | 0.46 | <0.001 | 6.9 | <0.001 | 1.9 |
| Females | | 681 | 2.08 | 0.42 | <0.001 | 7.8 | <0.001 | 3.8 | 2.08 | 0.42 | <0.001 | 7.8 | <0.001 | 3.8 |
| Australia | | 271 | 2.84 | 0.51 | <0.001 | 34.8 | <0.001 | 7.7 | 2.84 | 0.51 | <0.001 | 34.8 | <0.001 | 7.7 |
| Males | | 100 | 2.65 | 0.88 | 0.004 | 56.5 | <0.001 | 6.3 | 2.59 | 0.86 | 0.004 | 57.1 | <0.001 | 5.9 |
| Females | | 171 | 2.97 | 0.69 | <0.001 | 26.9 | <0.001 | 8.6 | 2.96 | 0.71 | <0.001 | 27.0 | <0.001 | 8.5 |
| The United States (Males) | | 1322 | 1.98 | 0.29 | <0.001 | 5.8 | <0.001 | 4.0 | 2.01 | 0.29 | <0.001 | 6.4 | <0.001 | 4.1 |
| Finland (Females) | | 428 | 2.50 | 0.54 | <0.001 | 9.4 | 0.002 | 6.2 | 2.51 | 0.54 | <0.001 | 9.6 | 0.003 | 6.2 |

Note: The results are from linear mixed-effects regression models. The model without BMI included zPGS HGS as a predictor and was adjusted for age at the first available HGS

measurement and 10 principal genetic components. The model with BMI was additionally adjusted for BMI. The analysis in full cohort was further adjusted for country. HGS was

standardized by sex. Family number was included as a random effect to account for relatedness. ΔR^2^ indicates the difference in the coefficient of determination (R^2^) between models with

and without PGS HGS. z, standardized for normal distribution; PGS, polygenic score; HGS, handgrip strength; BMI, body mass index; SE, standard error.

**eTable 3. Interactions Between zPGS HGS and Age**

| Country | N | β | SE | *P* |
| --- | --- | --- | --- | --- |
| All | 5103 | -0.016 | 0.014 | 0.258 |
| Males | 2858 | -0.006 | 0.021 | 0.779 |
| Females | 2245 | -0.038 | 0.020 | 0.058 |
| Denmark | 1870 | -0.048 | 0.020 | 0.019 |
| Males | 907 | -0.011 | 0.030 | 0.723 |
| Females | 963 | -0.076 | 0.026 | 0.004 |
| Sweden | 1210 | -0.041 | 0.033 | 0.218 |
| Males | 528 | -0.039 | 0.052 | 0.446 |
| Females | 682 | -0.048 | 0.043 | 0.269 |
| Australia | 271 | -0.112 | 0.095 | 0.241 |
| Males | 100 | -0.152 | 0.136 | 0.265 |
| Females | 171 | -0.121 | 0.117 | 0.304 |
| The United States (Males) | 1323 | -0.110 | 0.080 | 0.169 |
| Finland (Females) | 429 | 0.116 | 0.144 | 0.424 |

Note: The results are from linear mixed-effects regression models. The full model

included both the main effects of age at the first available HGS measurement and

zPGS HGS, as well as an interaction term (zPGS HGS × Age). Models were adjusted

for the first 10 genetic principal components. Analysis in the full cohort was

additionally adjusted for country. HGS was standardized by sex. Family number was

included as a random effect to account for relatedness. z, standardized for normal

distribution; PGS, polygenic score; HGS, handgrip strength; SE, standard error.

**eTable 4. Interactions Between zPGS HGS and Country**

| Variable | β | SE | *P* |
| --- | --- | --- | --- |
| Australia (reference) |  |  |  |
| Denmark | -1.317 | 0.609 | 0.031 |
| Sweden | -1.700 | 0.651 | 0.009 |
| The United States | -1.641 | 0.629 | 0.009 |
| Finland | -1.073 | 0.790 | 0.174 |

Note: The results are from linear mixed-effects regression models. The full model

included both the main effects of country and zPGS HGS, as well as an interaction term

(zPGS HGS × Country). Models were adjusted for the age at the first available HGS

measurement and the first 10 genetic principal components. HGS was standardized by sex.

Family number was included as a random effect to account for relatedness. z, standardized

for normal distribution; PGS, polygenic score; HGS, handgrip strength; SE, standard error.

**eFigure 6. Marginal Effects of PGS HGS Within Countries from Interaction	Models**

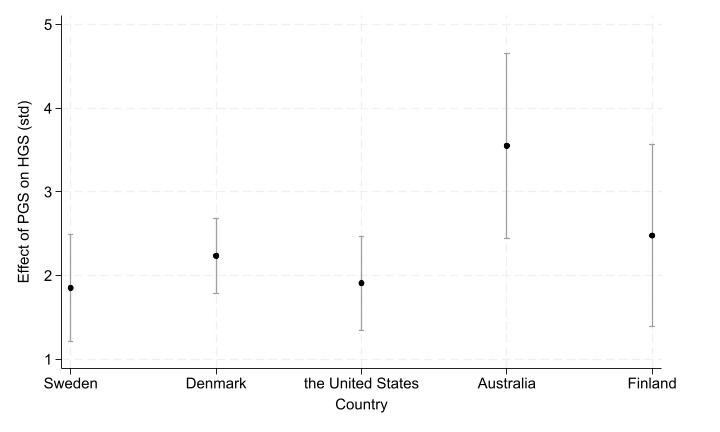

**eTable 5. Sample Characteristics of the Participants with Complete Case Data for HGS, BMI, and LTPA**

**Measurements (N = 4451)**

| Country/Cohort | N | Std HGS by sex^§^  mean (SD) | Age HGS  range (median) | BMI^†^  mean (SD) | Age BMI  range (median) | LTPA^‡^  mean (SD) | Age LTPA  range (median) |
| --- | --- | --- | --- | --- | --- | --- | --- |
| All | 4451 | -1.22 (9.21) | 48–96 (66) | 27.43 (4.77) | 48–98 (66) | 49.50 (10.03) | 45–96 (66) |
| Denmark | 1780 | -1.31 (9.22) | 53–96 (67) | 26.45 (4.15) | 53–98 (67) | 49.69 (10.08) | 54–96 (67) |
| *LSADT* | 326 | -1.56 (8.52) | 74–96 (79) | 24.60 (3.67) | 76–98 (80) | 50.52 (10.63) | 74–96 (78) |
| *MADT* | 1454 | -1.25 (9.37) | 53–85 (65) | 26.87 (4.14) | 53–85 (65) | 49.50 (9.95) | 54–85 (65) |
| Sweden | 984 | -0.82 (9.99) | 48–88 (72) | 26.30 (4.09) | 48–88 (72) | 49.19 (10.32) | 45–88 (71) |
| *SATSA* | 615 | -1.25 (10.00) | 48–88 (67) | 26.14 (4.14) | 48–88 (67) | 49.68 (10.14) | 45–88 (67) |
| *GENDER* | 369 | -0.11 (9.95) | 70–81 (74) | 26.56 (3.99) | 71–80 (74) | 48.39 (10.59) | 69–79 (73) |
| Australia *(OATS)* | 203 | 0.17 (9.56) | 70–94 (75) | 26.97 (4.47) | 70–94 (75) | 48.67 (9.29) | 65–90 (69) |
| U.S. *(VETSA)* | 1056 | -2.20 (7.86) | 53–67 (63) | 30.01 (5.38) | 57–67 (63) | 49.73 (9.71) | 57–67 (63) |
| Finland (*FITSA*) | 428 | -0.02 (10.00) | 63–76 (69) | 27.93 (4.73) | 63–76 (69) | 49.26 (10.23) | 63–76 (69) |

Note: § the	handgrip measurement and corresponding age closest to the physical activity assessment; †the BMI measurement and corresponding age closest to the handgrip

strength assessment; ‡ the first possible harmonized physical activity scores and corresponding age; HGS, handgrip strength; BMI, body mass index; LTPA; leisure-time physical

activity; SE, standard error.

**eFigure 7. Density Curves of LTPA among the IGEMS Participants and Stratified by Sex**

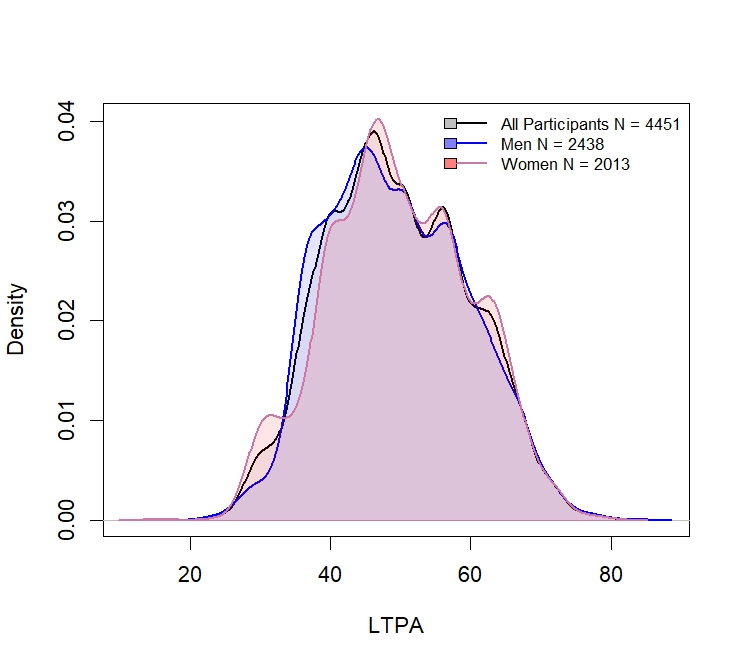

**eFigure 8. Density Curves of LTPA Stratified by Country**

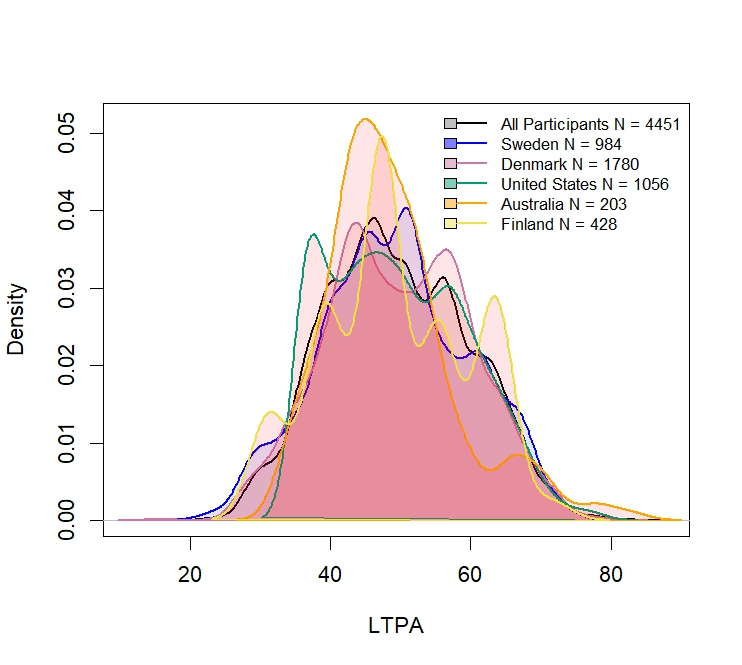

| Country | N | β | SE | *p* |
| --- | --- | --- | --- | --- |
| All | 4451 | -0.034 | 0.014 | 0.013 |
| Males | 2438 | -0.027 | 0.018 | 0.137 |
| Females | 2013 | -0.043 | 0.020 | 0.035 |
| Denmark | 1780 | -0.014 | 0.020 | 0.485 |
| Males | 872 | -0.003 | 0.026 | 0.907 |
| Females | 908 | -0.022 | 0.032 | 0.495 |
| Sweden | 984 | -0.034 | 0.030 | 0.251 |
| Males | 434 | -0.012 | 0.045 | 0.794 |
| Females | 550 | -0.042 | 0.037 | 0.260 |
| Australia | 203 | -0.182 | 0.065 | 0.006 |
| Males | 76 | -0.306 | 0.119 | 0.013 |
| Females | 127 | -0.240 | 0.081 | 0.004 |
| The United States (Males) | 1056 | -0.045 | 0.029 | 0.118 |
| Finland (Females) | 428 | -0.060 | 0.046 | 0.188 |

**eTable 6. Interactions Between zPGS HGS and LTPA**

Note: The results are from linear mixed-effects regression models. The full model included both the main

effects of zPGS HGS and LTPA, as well as an interaction term (zPGS HGS × LTPA). Models were adjusted

for age at HGS measurement (closest to the LTPA assessment), BMI (closest to HGS), and the first 10

genetic principal components. Analysis in the full cohort was additionally adjusted for country. HGS

was standardized by sex. Family number was included as a random effect to account for relatedness. z,

standardized for normal distribution; PGS, polygenic score; HGS, handgrip strength; LTPA, leisure-time

physical activity, BMI, body mass index; SE, standard error.

**eFigure 9. Interaction between PGS HGS and LTPA on sex-specific standardized HGS in Study II within the Australian cohort.**

.
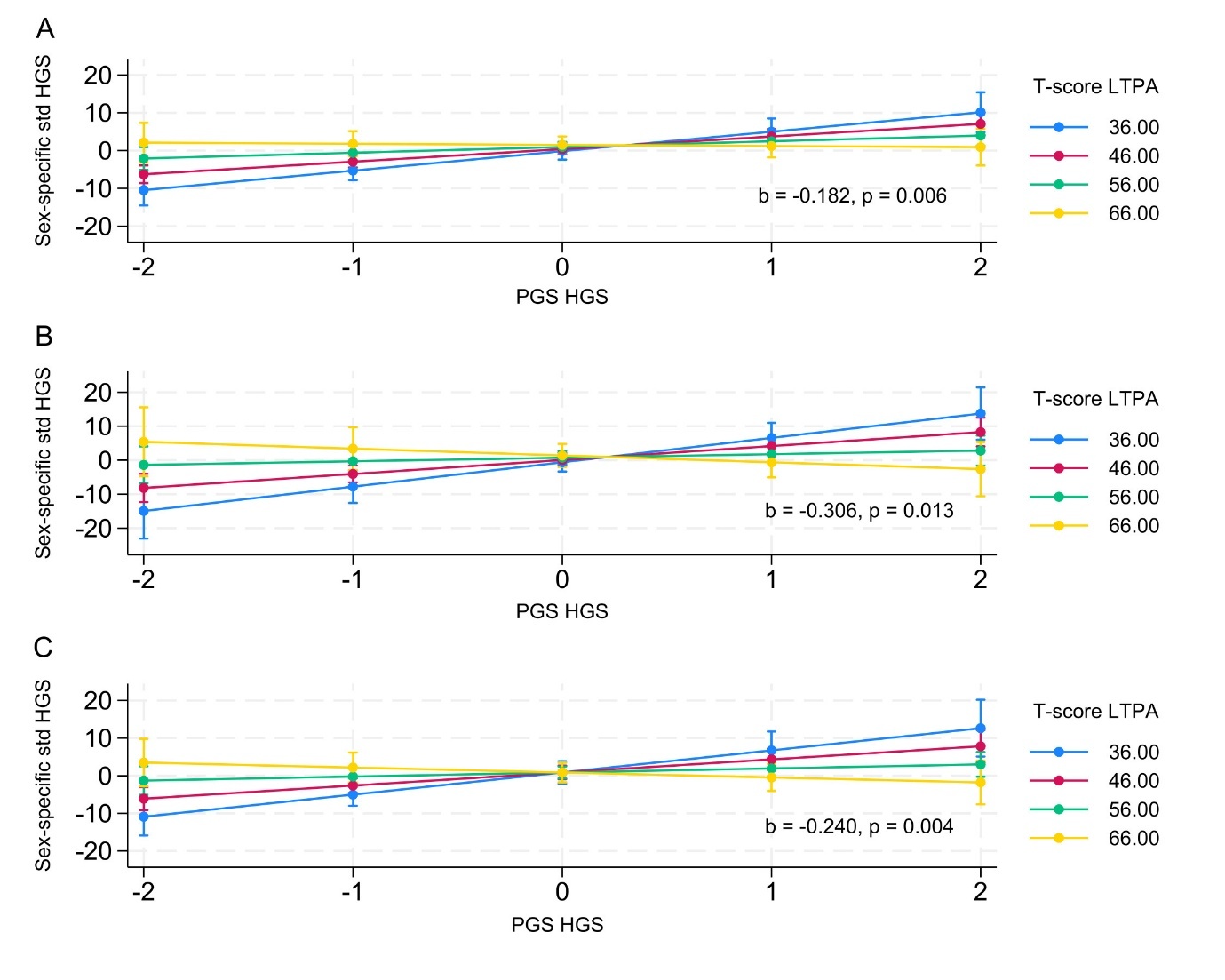

The figure shows that the positive effect of LTPA on HGS was stronger among individuals with a lower PGS HGS. The results are based on linear regression models incorporating the main effects of PGS HGS and LTPA, along with their interaction term (PGS HGS × LTPA). Models are adjusted for age at HGS measurement, BMI, and 10 principal genetic components, with family number included as a random factor. Both PGS HGS and LTPA were treated as continuous variables in the models. For illustrative purposes, the figure depicts T-score cut-off points, where the lowest value represents the 5th percentile, the highest value represents the 95th percentile, and intermediate values are spaced at 10-point intervals. Panels represent: (A) both sexes in the Australian cohort; (B) males; (C) females. Beta coefficients (b) and p-values indicate the strength and statistical significance of the PGS HGS × LTPA interaction term. The plotted lines represent estimated associations, and the error bars indicate 95% CIs

**eTable 7. Descriptive Characteristics of the Twin Pairs Included**

**in the Within-Twin Pair Analysis**

| Descriptives | All pairs (N=1982) | DZ pairs  (N=1124) | MZ pairs  (N=858) |
| --- | --- | --- | --- |
| Males (%) | 55 | 52 | 59 |
| Age | 66.56 (7.11) | 67.14 (7.07) | 65.47 (7.35) |
| HGS | -1.09 (9.11) | -0.51 (9.06) | -2.03 (8.85) |
| LTPA | 49.47 (10.04) | 49.29 (10.08) | 49.66 (9.93) |
| BMI | 27.47 (4.76) | 27.23 (4.60) | 27.78 (4.97) |

Note: Participants were restricted to twin pairs in which both individuals had complete data on HGS,

BMI, and LTPA. Values are presented as mean and standard deviation (SD), unless otherwise

stated. Age at the handgrip strength assessment corresponds to the LTPA assessment. DZ,

dizygotic; MZ, monozygotic; LTPA, leisure-time physical activity; HGS, handgrip strength; BMI, body

mass index.

**eTable 8. Mean and Standard Deviation of Within-Pair Differences**

|  | All (N=1982) | DZ pairs (N=1124) | MZ pairs (N=858) |
| --- | --- | --- | --- |
| Variable | Mean (SD) | Mean (SD) | Mean (SD) |
| Age | 0.23 (1.09) | 0.20 (0.92) | 0.26 (1.28) |
| HGS | 7.72 (6.59) | 8.49 (6.81) | 6.71 (6.14) |
| LTPA | 9.58 (7.94) | 10.26 (8.19) | 8.70 (7.52) |
| BMI | 3.52 (3.14) | 4.08 (3.41) | 2.78 (2.56) |

Note: Participants were restricted to twin pairs in which both individuals had complete data on HGS, BMI, and

LTPA. Age, LTPA, and BMI differences are based on absolute within-pair values. Values are presented as mean

and standard deviation (SD). Age at the handgrip strength assessment corresponds to the LTPA assessment.

DZ, dizygotic; MZ, monozygotic; LTPA, leisure-time physical activity; HGS, handgrip strength; BMI, body mass

index.

|  | β | SE | 95% CI | *P* |
| --- | --- | --- | --- | --- |
| All pairs (N=1982) | 0.059 | 0.018 | 0.023–0.094 | 0.001 |
| Dizygotic twin pairs (N=1124) | 0.090 | 0.025 | 0.042–0.139 | <0.001 |
| Monozygotic twin pairs (N=858) | 0.004 | 0.027 | -0.049–0.056 | 0.894 |
| By genetic liability to HGS^§^ |  |  |  |  |
| Low | 0.025 | 0.042 | -0.059–0.108 | 0.560 |
| Intermediate | 0.018 | 0.046 | -0.072–0.109 | 0.689 |
| High | -0.056 | 0.049 | -0.152–0.041 | 0.259 |

**eTable 9. The Association Between LTPA and HGS: Within-Twin Pair Analysis**

Note: The results are from the the fixed-effects within-pair regression models. Models are adjusted for age and BMI. ^§^Analysis restricted to monozygotic twin pairs stratified by PGS HGS tertiiles. LTPA, leisure-time physical activity; HGS, handgrip strength; PGS, polygenic score; CI, confidence interval. BMI, body mass index.

**eFigure 10. Marginal Effects of LTPA Within DZ Twin Pairs from Interaction Models**

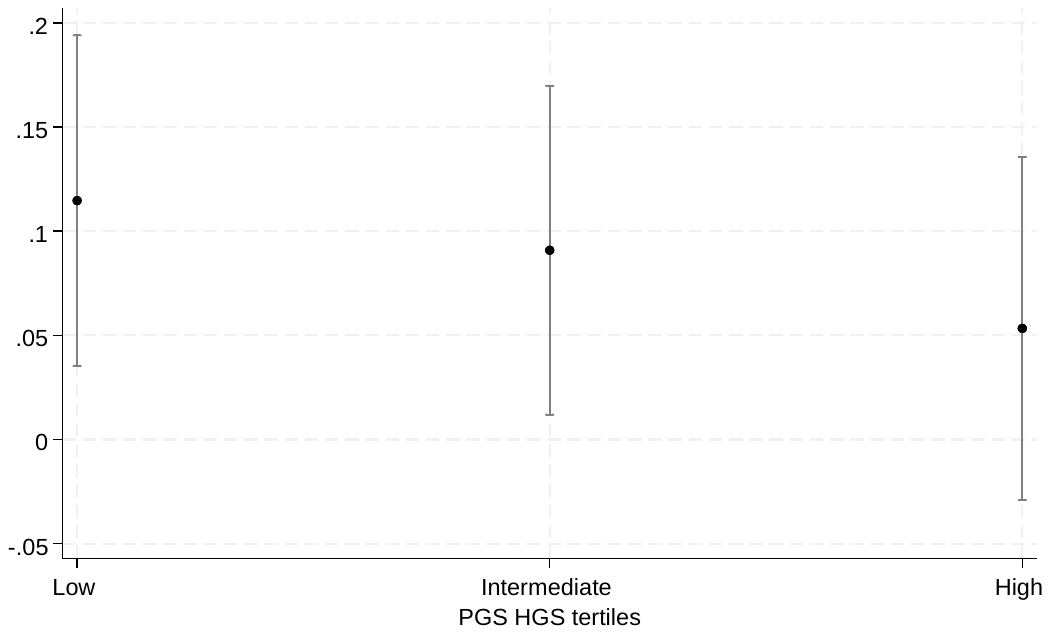

Note: Fixed-effect within-twin-pair model among DZ twins. Marginal effects estimated from interaction models (PGS HGS tertiles × LTPA). LTPA, leisure-time physical activity; DZ, dizygotic. HGS, handgrip strength; PGS, polygenic score.

**eFigure 11. The Stratified Regression Coefficients for MZ Twins by PGS HGS Tertiles**

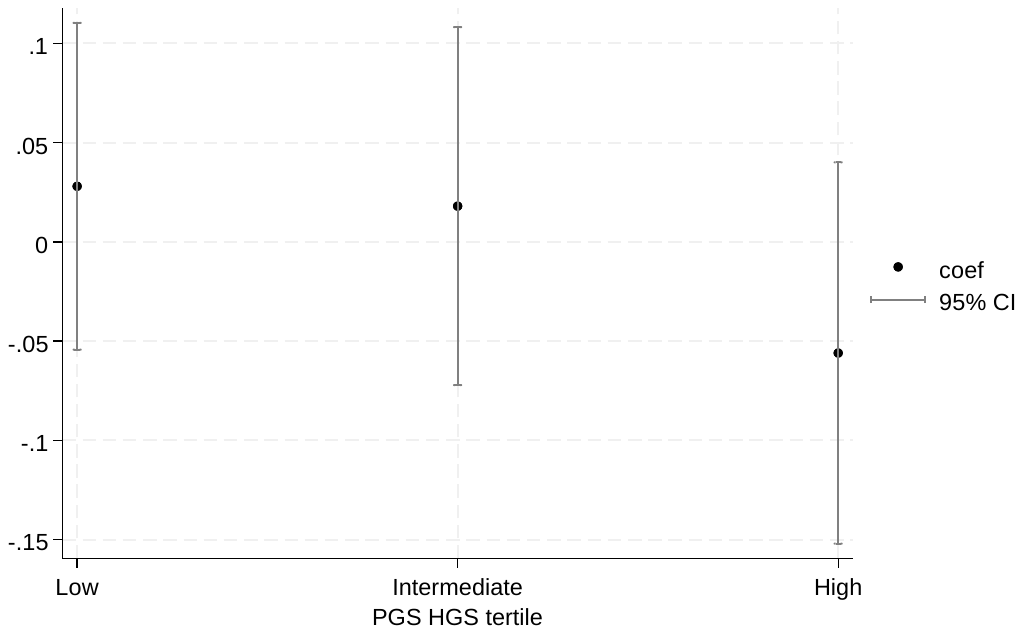

Note: Fixed-effect within-twin-pair model among MZ twins. Regression coefficients are from stratified models by PGS HGS tertiles. LTPA, leisure-time physical activity; MZ, monozygotic. HGS, handgrip strength; PGS, polygenic score; CI, confidence interval.

**Funding for the IGEMS Studies**

IGEMS is supported by the National Institutes of Health Grants No. R01 AG089666, R01 AG081248, R01 AG087486, and previously by R01 AG059329, R01 AG060470, RF1 AG058068, R01 AG037985 and R56 AG037985. SATSA was supported by grants R01 AG04563, R01 AG10175, the John D. and Catherine T. MacArthur Foundation Research Network on Successful Aging, the Swedish Council For Working Life and Social Research (FAS) (97:0147:1B, 2009-0795), and Swedish Research Council (825-2007-7460, 825-2009-6141). OCTO-Twin was supported by grant R01 AG08861. Gender was supported by the MacArthur Foundation Research Network on Successful Aging, The Axel and Margaret Ax:son Johnson’s Foundation, The Swedish Council for Social Research, and the Swedish Foundation for Health Care Sciences and Allergy Research. The Danish Twin Registry is supported by grants from The National Program for Research Infrastructure 2007 from the Danish Agency for Science and Innovation, the Velux Foundation and the US National Institute of Health (P01 AG08761). VETSA was supported by National Institute of Health grants NIA R01 AG018384, R01 AG018386, R01 AG022381, and R01 AG022982, and, in part, with resources of the VA San Diego Center of Excellence for Stress and Mental Health. The Cooperative Studies Program of the Office of Research & Development of the United States Department of Veterans Affairs has provided financial support for the development and maintenance of the Vietnam Era Twin (VET) Registry. Data collection and analyses in the Finnish Twin Cohort have been supported by ENGAGE – European Network for Genetic and Genomic Epidemiology, FP7-HEALTH-F4-2007, grant agreement number 201413, National Institute of Alcohol Abuse and Alcoholism (grants AA-12502, AA-00145, and AA-09203, the Academy of Finland Center of Excellence in Complex Disease Genetics (grant numbers: 213506, 129680),  and the Academy of Finland (grants 100499, 205585, 118555, 141054, 265240, 263278, 264146, 308248, and 312073). FITSA was supported by grants from the Academy of Finland (69818) and the Finnish Ministry of Education and Culture (120/722/2003). We acknowledge the contribution of the OATS research team (https://cheba.unsw.edu.au/project/older-australian-twins-study) to this study. The OATS study has been funded by a National Health & Medical Research Council (NHMRC) and Australian Research Council (ARC) Strategic Award Grant of the Ageing Well, Ageing Productively Program (ID No. 401162) and NHMRC Project Grants (ID 1045325 and 1085606). OATS participant recruitment was facilitated through Twins Research Australia, a national resource in part supported by a Centre for Research Excellence Grant (ID: 1079102), from the National Health and Medical Research Council. DNA from OATS participants was extracted by Genetic Repositories Australia, which was funded by the NHMRC Enabling Grant 401184 and genotyping was partly funded by a Commonwealth Scientific and Industrial Research Organisation Fl Collaboration Fund Grant.

We thank the participants for their time and generosity in contributing to this research. The content of this manuscript is solely the responsibility of the authors and does not necessarily represent the official views of the NIA/NIH, or the VA.

**Data availability for IGEMS Studies**

IGEMS data are not publicly available given the variety of data agreements and regulations governing the different studies and countries. However, many of the individual studies participating in IGEMS do have ways to access their data, some by direct request to the participating study, and many of the datasets may be accessed through National Archive of Computerized Data on Aging (NACDA). See https://doi.org/10.3886/ICPSR03843.v2 (SATSA), https://doi.org/10.3886/ICPSR25963.v2 (Study of Dementia in Swedish Twins), https://doi.org/10.3886/ICPSR02760.v19 (MIDUS), and https://www.icpsr.umich.edu/web/NACDA/studies/36234/versions/V6 (National Academy of Sciences-National Research Council Twin Registry). Two studies may be requested through Maelstrom: OCTO-Twin https://www.maelstrom-research.org/study/octo-twin and GENDER https://www.maelstrom-research.org/study/gender. For access to data from the Danish Twin Registry, see https://www.sdu.dk/en/om_sdu/institutter_centre/ist_sundhedstjenesteforsk/centre/dtr/researcher. To request OATS data please contact the CHeBA Research Bank via email on for a current application form. For VETSA data, see instructions to researchers: https://medschool.ucsd.edu/som/psychiatry/research/VETSA/Researchers/Pages/default.aspx, and for FITSA data (https://thl.fi/en/research‐and‐development/thl‐biobank/for‐researchers/application‐process).
